# Prospective Quantification of Tricuspid Regurgitation with Echocardiography vs 4D Flow Cardiac Magnetic Resonance

**DOI:** 10.1101/2024.10.11.24315310

**Authors:** Agata Sularz, Ahmed S. Negm, Alejandra Chavez-Ponce, Ahmed El Shaer, Chia-Hao Liu, Jared Bird, Jae Oh, Sorin Pislaru, Jeremy D. Collins, Mohamad Alkhouli

## Abstract

**Introduction:** Cardiac magnetic resonance (CMR) is a valuable tool in the assessment of valvular disease. However, its utilization in tricuspid regurgitation (TR) evaluation has been limited. We sought to compare TR grading with 4D-CMR and transthoracic echocardiography (TTE).

**Methods:** We prospectively recruited patients with ≥moderate TR on TTE to undergo multiparametric CMR with integrated cardiac function and 4D flow assessments using a 1.5-T scanner (Siemens Somatom Aera, Erlangen, Germany). Patients with other severe valvulopathy, end-stage renal disease, or pacemakers were excluded. TR was graded severe on CMR when TR volume ≥45 mL, and/or TR fraction ≥50%. The weighted kappa test was used to assess the agreement in overall TR grading on TTE and MR.

**Results:** Fifty-two patients were enrolled (mean age 78.5±7.6 years, 53.8% men). The median interval between CMR and TTE was 2 days (IQR=1-37). The agreement between TTE and CMR-derived TR volume was fair (kappa = 0.28, 95%CI 0.13-0.45), with only 10 of 31 patients (32%) with ≥severe TR on TTE meeting severe TR volume criterion on CMR (TR volume ≥45 ml). There was no agreement between TTE and CMR-derived TR fraction (kappa=0.04, 95%CI 0.13-0.46) with only 3 of 31 patients (13%) with ≥severe TR on TTE meeting severe TR criterion on CMR (TR fraction ≥50%).

**Conclusion:** Grading of TR was frequently discordant between TEE and 4D-MRI. Further studies are needed to elucidate the clinical impact of concordant/discordant TR grading on multi-modality imaging.

---

Tricuspid regurgitation (TR) is common and associated with right ventricular (RV) dysfunction and all-cause mortality^1^. Tricuspid valve surgery is typically reserved for symptomatic patients or those with progressive RV failure, and is associated with high perioperative morbidity and mortality^2,3^. However, surgical outcomes have been attributed to inadequate understanding of the natural progression of TR, its classification, and the optimal timing of intervention^4–8^. The advent of transcatheter interventions for TR have recently fueled a growing interest in bridging these knowledge gaps. In this context, many recent studies explored various methods for grading TR on echocardiography ^5,9–11^. These studies emphasized the challenges in accurate quantification of TR, suggesting the need to explore the role of other imaging modalities in grading TR.

Cardiac magnetic resonance (CMR) is increasingly recognized as a useful tool for the assessment of valvular heart disease^12–15^. Nevertheless, the accuracy of CMR in estimating TR can be affected by the exaggerated motion of the tricuspid annulus^12^, difficulty with breath-holding, and arrhythmia. Newer volumetric techniques such as four-dimensional (4D) flow CMR can more accurately assess flow across the valve and track the position of the tricuspid annulus over time^16^. In addition, 4D flow CMR enables single-volume acquisition of the entire heart and does not require specialized knowledge of cardiac anatomy and imaging planes. This allows TR jets to be directly quantified with a single measurement eliminating the need for ventricular endocardial contouring and increasing reporting efficiency^16,17^. Few studies assessed the agreement between the TTE and CMR in quanitifying TR^16,18,19^, similar to what has been reported on the quantification of mitral and aortic regurgitation^20–24^. However, except for in the context of congenital heart disease, these comparative studies did not include 4D flow CMR assessment. We compared TR grading by TTE vs multiparametric CMR with integrated 4D flow analysis in patients with ≥moderate regurgitation at TTE. Additionally, we investigated the correlation between CMR-derived TR metrics and RV volumes.

## Methods

### Study Population

We prospectively enrolled patients referred to the structural heart disease clinic at Mayo Clinic, MN between January 2022 and December 2023. Patients with moderate or greater TR on the most recent TTE were included (within 45 days without apparent change in volume status). We excluded patients with other significant valvulopathy (>moderate regurgitation, and/or stenosis), end-stage renal disease, permanent pacemakers, claustrophobia, and pregnant women. All patients provided written consent. This prospective study was registered on ClinicalTrials.gov (NCT05006443).

### CMR acquisition

Comprehensive CMR scans were acquired on a 1.5-T MRI scanner (MAGNETOM Aera, Siemens Healthineers, Erlangen, Germany). Two cine sequences were acquired for each subject: the standard breath-hold segmented cine (Seg Cine), and the free-breathing compressed-sensing sequence with motion correction (FB-MOCO) (**Supplement Table 1**). 2D phase contrast imaging (2D flow) was obtained through the main pulmonary artery, aortic root at the sinotubular junction (STJ), through the open leaflets of the mitral valve in diastole, and through the tricuspid annulus in diastole. 4D flow MRI was performed post-contrast with whole heart coverage. Acquisition parameters are presented in the supplementary material.

### CMR analysis

We used Circle Cvi42 version 5.13.5 (Circle Cardiovascular Imaging Inc., Calgary, Canada), an AI-assisted segmentation software, for augmented manual postprocessing. An MD researcher with two years of postprocessing training performed all aspects of CMR analysis, including the SAX cine stack biventricular, 2D flow and 4D flow analysis (**Supplemental Figures 1-7**). A cardiovascular radiologist with 13 years experience validated all measurements. We assessed biventricular size and systolic function using Simpson’s method on the SAX cine imaging. Segmented cine images were used for biventricular systolic analysis if of diagnostic quality; otherwise biventricular systolic function was derived from FB-MOCO cine images. Valvular function was assessed on 2D and 4D flow contrast phases. Regurgitant volume and fraction were estimated using total forward and backward volumes (2D and 4D flow contrast phase only). We then graded TR severity according to thresholds described elsewhere: ^25^

I. Based on TR volume: mild <30 mL; moderate = 30 to 44 mL; severe ≥45 mL by CMR.
II. Based on TR fraction: mild <30%; moderate = 30% to 49%; severe ≥50%^25^.

In a separate analysis, we graded TR according to TR volume indexed by Body Surface Area (BSA).

### Echocardiography

All patients underwent standard-of-care TTE with commercially available systems. Standard views were acquired. At our institution, TR is graded as trivial, mild, mild-moderate, moderate, moderate-severe or severe based on the integration of several parameters according to the American Society of Echocardiography guidelines^26^. Those parameters include valve morphology, color Doppler TR jet, continuous-wave Doppler signal of the regurgitant jet, vena contracta width, proximal flow convergence zone, systolic hepatic vein flow reversal, inferior vena cava maximal diameter and tricuspid inflow velocities. The specific parameters used to grade TR were left to the discretion of the operator, and additional images were obtained as needed. All echos were subsequently reviewed and adjudicated by a dedicated independent core laboratory.

### Statistical analysis

Continuous data are expressed as means ± standard deviation (SD). Categorical data are presented as counts and frequencies (%). We assessed agreement in non-severe vs severe TR classification between the two imaging modalities using the weighted kappa test with quadratic weight. The calculated kappa coefficients were interpreted as: 0–0.20 low, 0.20–0.40 fair, 0.40–0.60 moderate, 0.60–0.80 good, and >0.80 excellent. Additionally, we performed the McNemar test to assess for significant differences in the proportion of observations classified as severe vs non-severe TR. We used the Kruskal–Wallis test to compare TTE-based TR grades in terms of TR volume, TR fraction, and RV end-diastolic volume (RVEDV) indexed by BSA (details in the **Supplement Table 2**). For pairwise comparisons, we used the Dunn test with Bonferroni correction. Finally, we used the Spearman correlation coefficient (rho) to assess the correlation between TR volume and TR fraction, indexed TR volume and TR fraction, TR volume and indexed RVEDV, indexed TR volume and indexed RVEDV, and TR fraction and indexed RVEDV. *P* value < 0.05 was considered statistically significant. All statistical analyses were performed using the R studio software (Version 2023.09.1+494, Posit Software, PBC).

## Results

### Patient selection and baseline characteristics

Fifty-two patients were enrolled (**Figure 1**). 2D flow and 4D flow CMR data were interpretable in 46/52 (88%) and 49/52 (94%) patients, respectively. Mean age was 78.5±7.6 and 28/52 (53.8%) were men (**Table 1**). The median interval between CMR and TTE was 2 days (IQR = 1-37). Seven patients had primary TR; the remainder (45/52) had non-pacemaker related secondary TR. RV dilatation and impaired RV systolic function were present in 45/52 (87%) and 29/52 (56%) of patients (**Table 1, Supplemental Table 4**). **Figure 2**. illustrates the distribution of TR grading by TTE, CMR-derived TR volume and TR fraction. **Table 1**. and **Supplemental Tables 5-7** summarize the CMR results.

**Figure 1.**
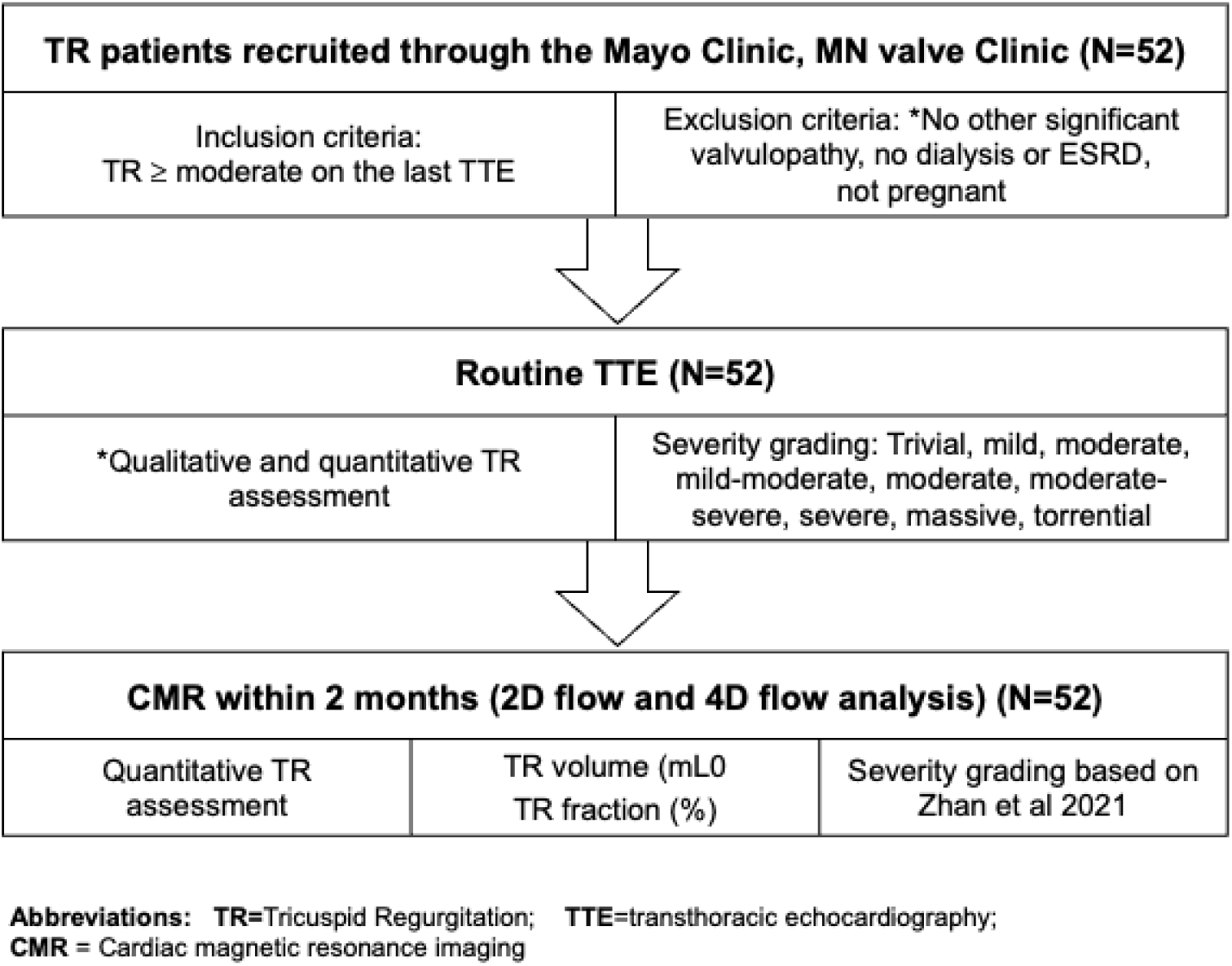
Study flow chart.

**Figure 2.**
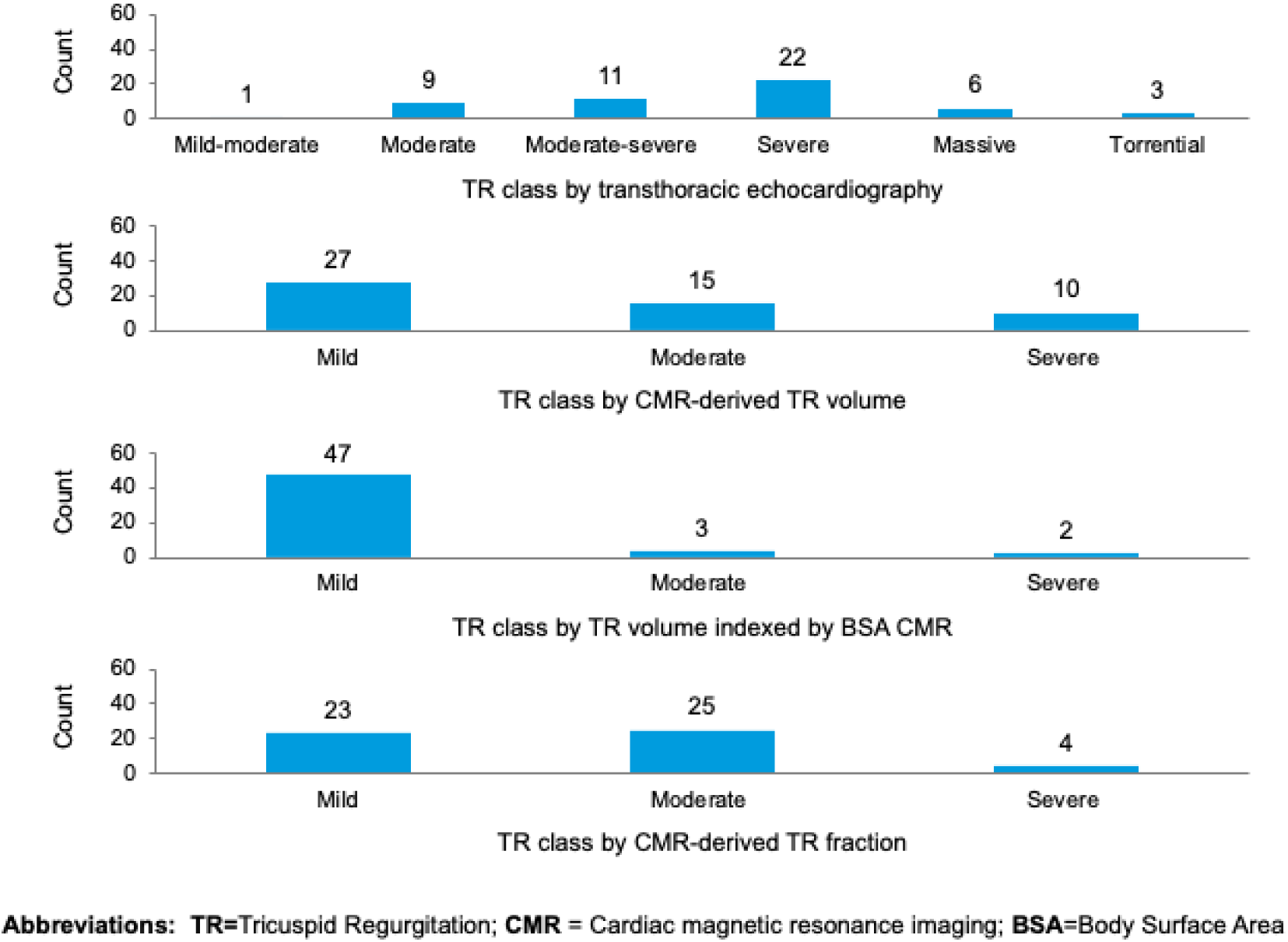
The distribution of tricuspid regurgitation (TR) grading by transthoracic echocardiography, cardiac magnetic resonance (CMR)-derived TR volume and TR fraction.

**Table 1.**
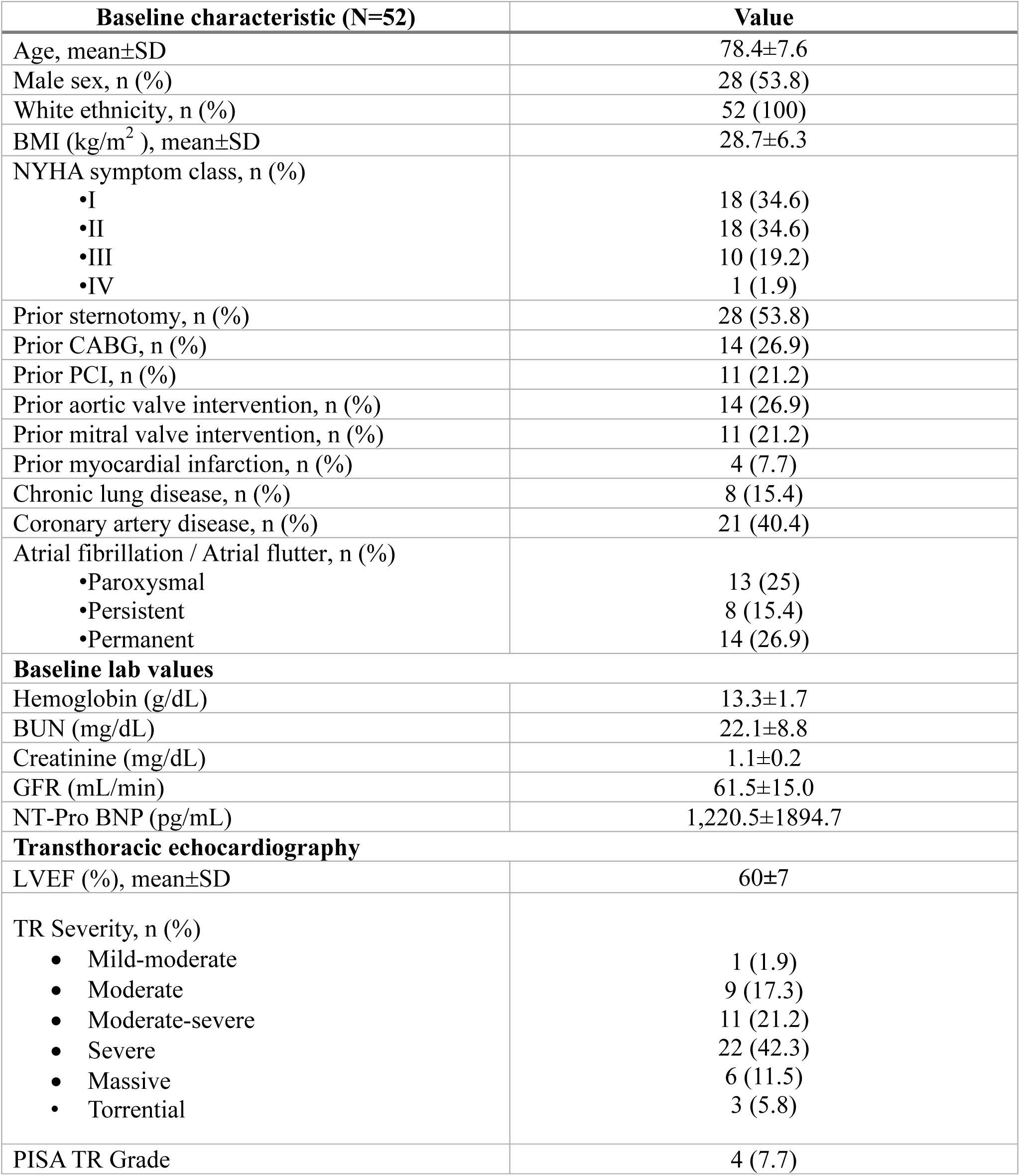
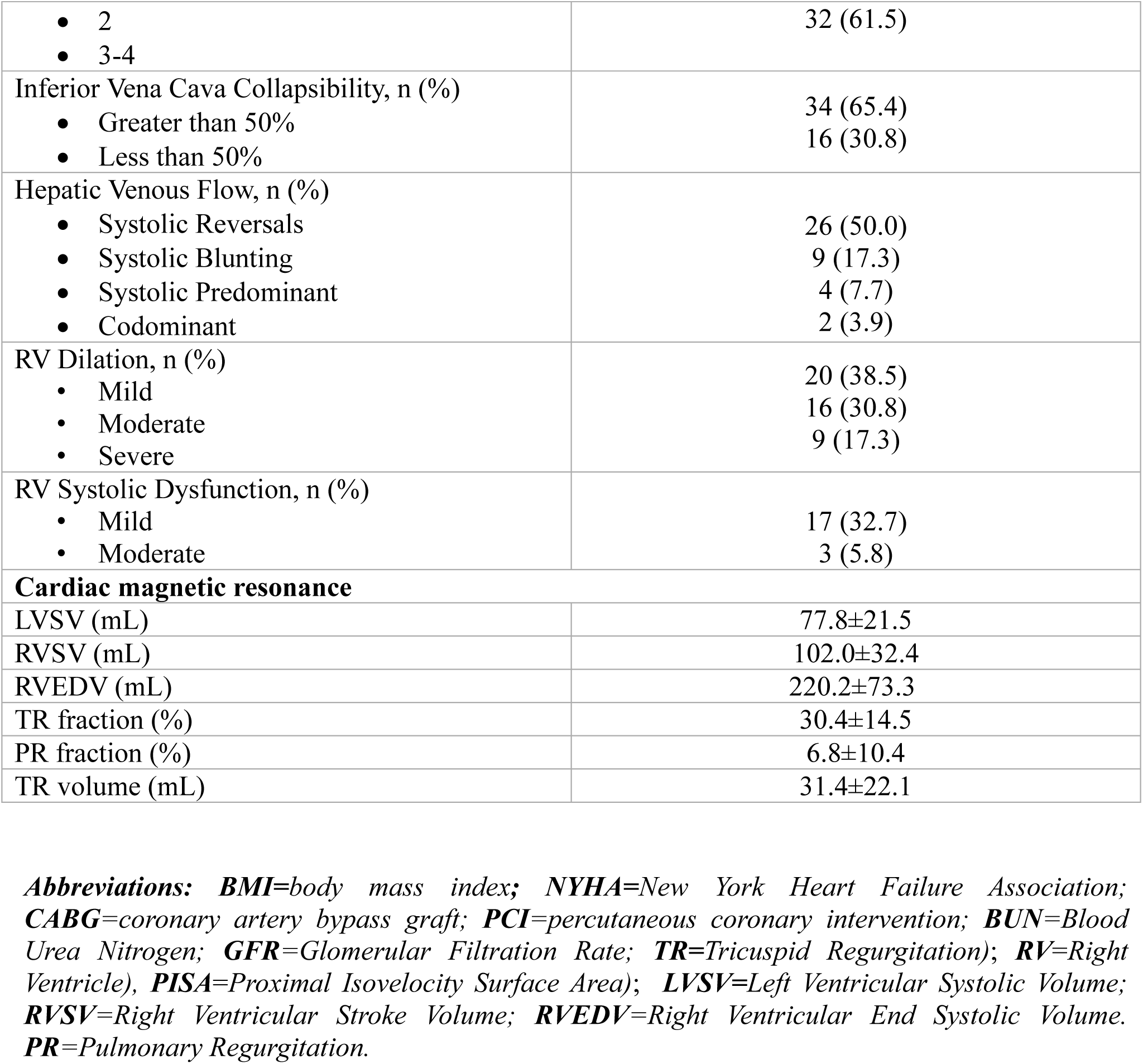
Baseline characteristics, laboratory and imaging results.

| **Baseline characteristic (N=52)** | **Value** |
| --- | --- |
| Age, mean±SD | 78.4±7.6 |
| Male sex, n (%) | 28 (53.8) |
| White ethnicity, n (%) | 52 (100) |
| BMI (kg/m<sup>2</sup>), mean±SD | 28.7±6.3 |
| NYHA symptom class, n (%) |  |
| •I | 18 (34.6) |
| •II | 18 (34.6) |
| •III | 10 (19.2) |
| •IV | 1 (1.9) |
| Prior sternotomy, n (%) | 28 (53.8) |
| Prior CABG, n (%) | 14 (26.9) |
| Prior PCI, n (%) | 11 (21.2) |
| Prior aortic valve intervention, n (%) | 14 (26.9) |
| Prior mitral valve intervention, n (%) | 11 (21.2) |
| Prior myocardial infarction, n (%) | 4 (7.7) |
| Chronic lung disease, n (%) | 8 (15.4) |
| Coronary artery disease, n (%) | 21 (40.4) |
| Atrial fibrillation / Atrial flutter, n (%) |  |
| •Paroxysmal | 13 (25) |
| •Persistent | 8 (15.4) |
| •Permanent | 14 (26.9) |
| **Baseline lab values** |  |
| Hemoglobin (g/dL) | 13.3±1.7 |
| BUN (mg/dL) | 22.1±8.8 |
| Creatinine (mg/dL) | 1.1±0.2 |
| GFR (mL/min) | 61.5±15.0 |
| NT-Pro BNP (pg/mL) | 1,220.5±1894.7 |
| **Transthoracic echocardiography** |  |
| LVEF (%), mean±SD | 60±7 |
| TR Severity, n (%) |  |
| • Mild-moderate | 1 (1.9) |
| • Moderate | 9 (17.3) |
| • Moderate-severe | 11 (21.2) |
| • Severe | 22 (42.3) |
| • Massive | 6 (11.5) |
| • Torrential | 3 (5.8) |
| PISA TR Grade | 4 (7.7) |
| • 2<br>• 3-4 | 32 (61.5) |
| Inferior Vena Cava Collapsibility, n (%)<br>• Greater than 50%<br>• Less than 50% | 34 (65.4)<br>16 (30.8) |
| Hepatic Venous Flow, n (%)<br>• Systolic Reversals<br>• Systolic Blunting<br>• Systolic Predominant<br>• Codominant | 26 (50.0)<br>9 (17.3)<br>4 (7.7)<br>2 (3.9) |
| RV Dilation, n (%)<br>• Mild<br>• Moderate<br>• Severe | 20 (38.5)<br>16 (30.8)<br>9 (17.3) |
| RV Systolic Dysfunction, n (%)<br>• Mild<br>• Moderate | 17 (32.7)<br>3 (5.8) |
| **Cardiac magnetic resonance** |  |
| LVSV (mL) | 77.8±21.5 |
| RVSV (mL) | 102.0±32.4 |
| RVEDV (mL) | 220.2±73.3 |
| TR fraction (%) | 30.4±14.5 |
| PR fraction (%) | 6.8±10.4 |
| TR volume (mL) | 31.4±22.1 |

**Abbreviations:** *BMI*=body mass index; *NYHA*=New York Heart Failure Association; *CABG*=coronary artery bypass graft; *PCI*=percutaneous coronary intervention; *BUN*=Blood Urea Nitrogen; *GFR*=Glomerular Filtration Rate; *TR*=Tricuspid Regurgitation); *RV*=Right Ventricle), *PISA*=Proximal Isovelocity Surface Area); *LVSV*=Left Ventricular Systolic Volume; *RVSV*=Right Ventricular Stroke Volume; *RVEDV*=Right Ventricular End Systolic Volume. *PR*=Pulmonary Regurgitation.

### Comparison of TR severity between TTE and CMR-derived TR volume

The agreement between TTE and CMR-derived TR volume in severe or higher vs non-severe TR classification was fair (kappa = 0.28, 95% confidence interval (CI) 0.13-0.45). There was complete agreement in 31 (60%) cases (**Table 2**). Only 10 of 31 patients (32%) with severe or higher TR on TTE had severe TR on CMR (TR volume ≥45 mL). The McNemar test result was consistent and indicated a statistically significant difference in classification performance between the two modalities (*p*<0.001).

**Table. 2.**
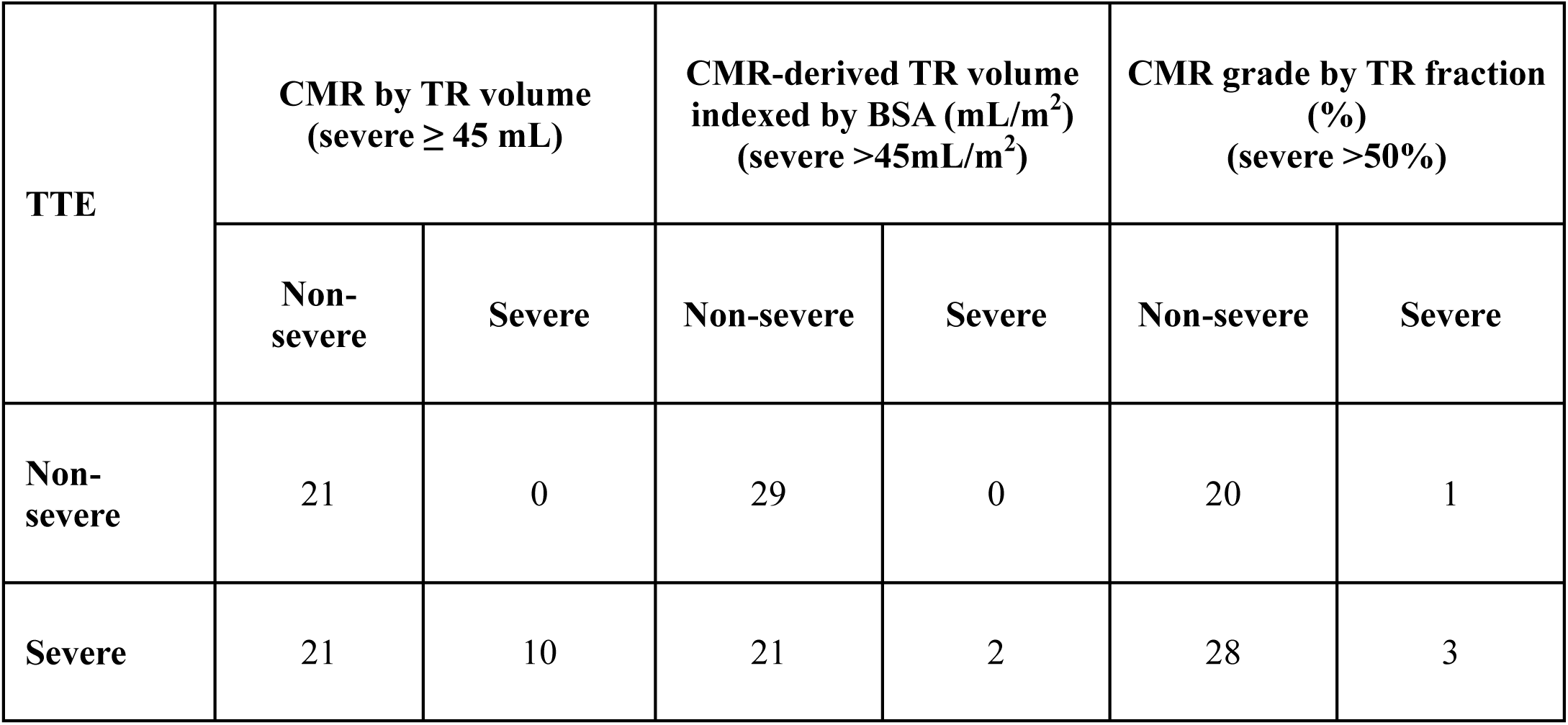
TR grade by transthoracic echocardiography (TTE) vs CMR-derived severity grading. All observations presented as counts.

### Comparison of TR severity between TTE and CMR-derived TR volume indexed by BSA

The agreement between severe vs non-severe TR classification by TTE and CMR (TR volume indexed by BSA mL/m^2^) was low (kappa 0.096, 95%CI 0.0-0.25). Only 2 of 23 patients (9%) with severe TR on TTE had severe TR on CMR (indexed TR volume ≥ 45 mL/m^2^) (**Table 2**). McNemar test result was consistent and indicated a statistically significant difference in classification performance between the two modalities (**p<0.001**).

### Comparison of TR severity between TTE and CMR-derived TR fraction

The agreement between TTE and CMR-derived by TR fraction was low (kappa 0.040, 95%CI 0.13-0.46). Only 3 of 31 patients (9.6%) with severe or greater TR on TTE had severe TR on CMR (TR fraction ≥50%) (**Table 2**). The McNemar test result was consistent and indicated a statistically significant difference in classification performance between the two modalities (*p* < 0.001).

### Comparison of CMR-derived TR volume, fraction and indexed RVEDV with TTE grade

TTE-based TR grades were significantly different in terms of CMR-derived TR volume (*p* < 0.001) (**Figure 3**). In pairwise comparisons, the severe or greater vs moderate and severe or greater vs moderate or less TR groups were significantly different (*p*=0.021, *p*<0.001). TR volume indexed by BSA (mL/m^2^) was significantly different between TTE-based grades (*p*<0.001). In pairwise comparisons, moderate vs severe and moderate-severe vs severe grades were significantly different (*p*<0.001 and p=0.007, respectively). TR fraction was also significantly different between groups (*p*<0.001). In pairwise comparisons, only the moderate vs severe or greater TR grades were significantly different (*p*<0.001). TTE grades were also significantly different in indexed RVEDV (mL/m^2^) (*p*=0.040). In pairwise comparisons, none of the groups were significantly different from each other (*p*>0.050).

**Figure 3.**
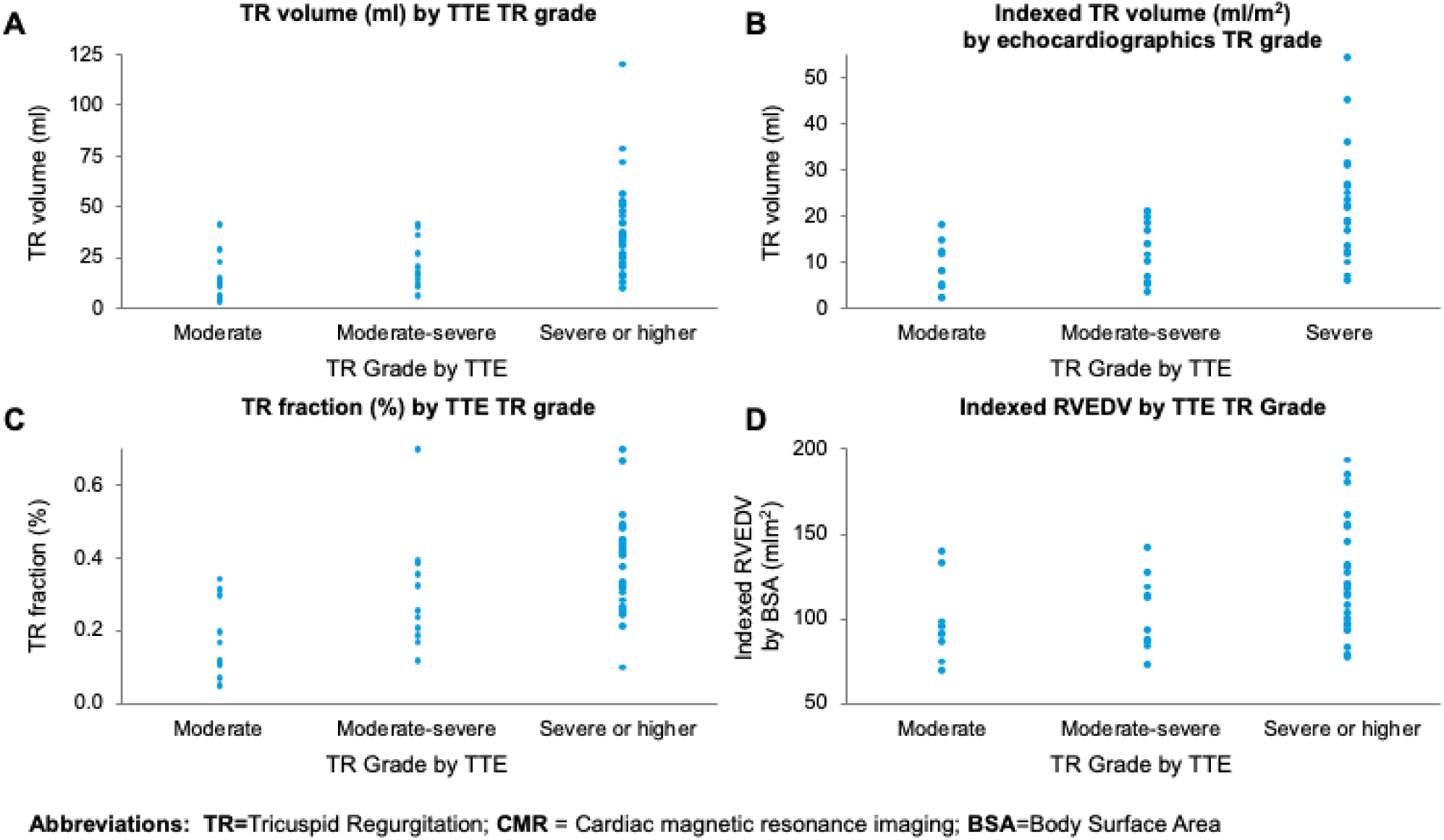
The distribution of (A) tricuspid regurgitant (TR) volume; (B) Indexed TR volume indexed; (C) TR fraction; (D) indexed Right Ventricular End-Diastolic Volume (RVEDV) Body Surface Area in patients with moderate or less, moderate-severe, severe or higher TR grades according to transthoracic echocardiography (TTE).

### Comparison of TR volume and TR fraction in the assessment of TR severity

There was a strong correlation between TR volume and TR fraction (rho=0.805, 95% CI= 0.372 - 0.923, *p*<0.001) (**Figure 4**). The correlation between TR volume and indexed RVEDV was moderate to strong (rho=0.60, 95%CI 0.38-0.74, *p*<0.001). The correlation between TR volume indexed by BSA (mL/m^2^) and indexed RVEDV was also moderate to strong (rho=0.58, 95%CI 0.37-0.73, *p*<0.001). The correlation between TR fraction and indexed RVEDV was only moderate (rho=0.37, 95%CI 0.09-0.59), *p*=0.008).

**Figure 4.**
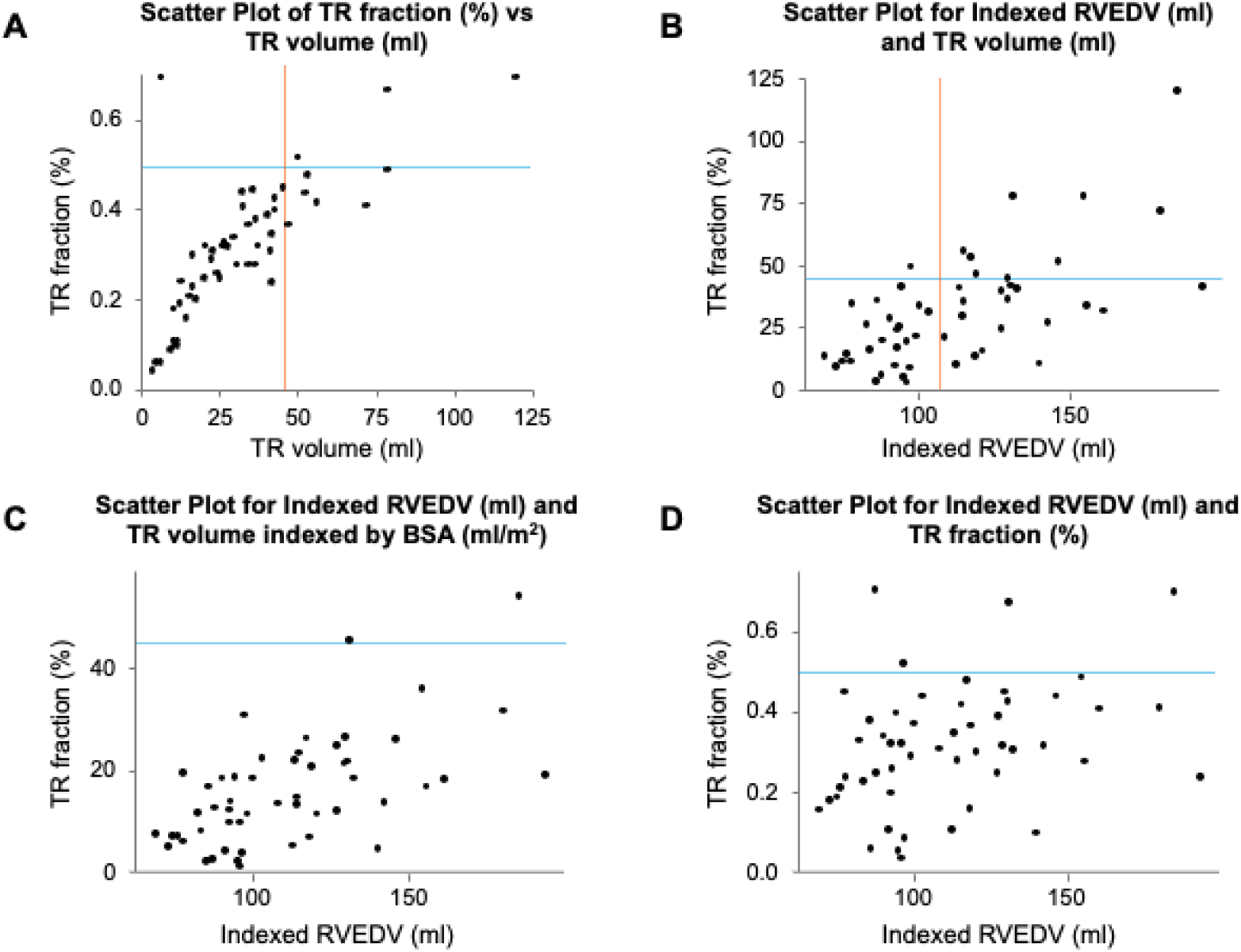
Correlation between (A) tricuspid regurgitant (TR) volume (mL) and TR fraction (%); (B) right ventricular end-diastolic volume (RVEDV) indexed by BSA (mL/m^2^) and TR volume; (C) indexed RVEDV and TR volume; (D) indexed RVEDV and TR fraction in patients with moderate or less, moderate-severe, severe or higher TR grades according to transthoracic echocardiography (TTE).

## Discussion

This is the first analysis of TR classification by TTE versus multiparametric CMR using integrated 4D flow analysis in patients with moderate or greater degrees of TR on TTE. We assessed agreement between modalities using established CMR thresholds^25^. We showed that there was limited agreement between the modalities (**Central Illustration**). For instance, 61% of patients with severe TR on TTE were classified as non-severe on CMR (as defined by TR volume <45 mL). When TR fraction was used to grade TR on CMR, 87% of patients with severe TR on TTE were classified as non-severe TR on CMR (TR fraction < 50%).

Assessment of TR with 4D flow CMR has been shown to be more accurate than traditional CMR ^17^. The increased accuracy stems from the ability of 4D flow to capture complex blood flow patterns in three dimensions over time. This enables a more comprehensive analysis of flow dynamics, especially in the presence of multiple TR jets. Moreover, the imaging for 4D flow analysis can be obtained in a single volume acquisition, which shortens scanning times and reduces the need for breath-holding and image post-processing^17^. 4D flow parameters have been validated against 2D flow parameters and 4D flow imaging has been previously used to assess TR in patients with complex congenital heart disease^27^.

Few studies have directly compared TR grading between TTE and CMR, and those leveraging 4D CMR included only patients with congential heart disease. Driessen et al measured 4D flow CMR-derived TR volume and fraction in 21 patients with congenital heart disease and pulmonary hypertension, 12 of whom had moderate or higher TR. Similar to our results, 75% of their patients were classified differently by at least one grade^16^. In a larger study, Zhan et al. compared TR grading by TTE and SAX CMR in 337 patients with functional TR. They assessed the diagnostic performance of several TTE parameters against CMR-derived TR volume to design a new hierarchical TTE algorithm based on the best-performing parameters. They then compared TR severity grading by the American Society of Echocardiography (ASE)-directed^26^ and hierarchical echocardiographic algorithms vs CMR. The hierarchical approach performed marginally better than the ASO-directed approach (65% vs 69% agreement with CMR-based TR grade). In most discordant cases, CMR suggested a less severe TR grade than TTE^18^. A similar trend was found by Medvedovsky et al in a study of 61 patients with isolated TR^19^. Our results are consistent with these reports; regardless of the parameter used to grade TR (volume or fraction), CMR-based grades were on average lower than TTE-based grades.

Unlike with TTE, there are no universal thresholds for TR grading in CMR; nevertheless, several grading systems have been proposed. Few studies used echocardiographic TR volume thresholds^19,26,28^, or leveraged similar CMR-derived thresholds that are used in the assessment of pulmonic^16,29^ or mitral valve regurgitation^30^. Recently, CMR-specific thresholds for TR volume and fraction have been derived from all-cause mortality outcome data in prospective cohort studies of patients with isolated severe TR^31^, functional TR^14,25^, and heart failure with reduced ejection fraction^32^. In our study, we used thresholds proposed by Zhan et al., the largest TR cohort studied thus far. After adjusting for clinical and imaging variables, including RV function, both TR fraction and TR volume were associated with increased mortality. Patients in the highest-risk group of TR volume ≥45 mL or TR fraction ≥50% had the worst prognosis. A summary of CMR thresholds for TR grading proposed in selected prospective studies of TR patients can be found in Table 3.

**Table 3.** Cardiac MRI (CMR) thresholds for TR grading proposed in selected prospective studies of tricuspid regurgitation (TR) patients. Abbreviations: HR: hazard ratio; aHR: adjusted hazard ratio; HFrEF: heart failure with reduced ejection fraction; HF: heart failure.

| Author | Year | Study type | Patient Population | Primary end point | N | Follow-up | CMR criteria for TR volume (mL)<br>(with aHR) | CMR criteria for TR fraction (%)<br>(with aHR) | CMR details |
| --- | --- | --- | --- | --- | --- | --- | --- | --- | --- |
| Zhan et al. | 2020 | Prospective cohort | Functional TR | **5-year** All cause mortality | 547 | Median 2.6 years | Low: <30 (reference)<br>Intermediate: 30 to 44 (1.46)<br>High: <sup>3</sup>45 (2.26) | Low: <30 (reference)<br>Intermediate: 30 to 49 (1.21)<br>High: <sup>3</sup>50 (2.60) | steady-state free-precession |
| Wang et al. | 2021 | Prospective cohort | Isolated severe TR | **10-year** All cause mortality | 262 | Mean 2.5 years | Low: <30<br>High: <sup>3</sup>30 | Minimal: <25<br>Low: 26-36<br>Intermediate: | steady-state free-precession |
|  |  |  |  |  |  |  | HR per 5 mL increase 1.05 (1.02-1.08) | 37 to 49<br>High: <sup>3</sup>50<br>HR per 5% increase 1.13 (1.06-1.21) |  |
| Seo et al. | 2023 | Prospective cohort | Patients with HFrEF, | **3-year:** A composite of all-cause death and hospitalization for HF | 262 | Median 921 days | Low: 10 (reference)<br>Intermediate: 10–30 (2.11 (1.15–3.89))<br>High: > 30 (7.16 (3.33–15.41)) | Low: ≤15 (reference)<br>Intermediate: 16–30 (2.26 (1.14–4.51))<br>High: >30 (4.23 (2.22–8.04)) | steady-state free-precession |
| Hinojar | 2021 | Prospective cohort | Patients with significant TR | **5-year:** A composite of hospital admission due to right heart failure and cardiovascular | 75 | Median 3 years | Low: <42 (reference)<br>High: <sup>3</sup>42 (HR (2.89))<br>TR volume per 1 mL | Low: <40 (reference)<br>High: <sup>3</sup>40 (HR 4.04)<br>TR fraction per 1 % aHR: | steady-state free-precession |
|  |  |  |  | mortality |  |  | aHR: 1.06<br>(1.03–1.08) | 1.02 (1.01–<br>1.03) |  |

Although TR volume and TR fraction correlate strongly, they are distinct parameters. Both parameters are a measure of the regurgitant volume, but only the regurgitant fraction takes into account the total volume of blood ejected by the ventricle, adjusting for systemic flow. In functional TR, which frequently occurs in a low-flow state, a relatively small TR volume can yield a large TR fraction. In a population with varying flow states, TR volume may not be as useful as TR fraction in accounting for a dilated RV. As an alternative, we decided to index TR volume by BSA. This allows normalization of TR volume, adjusting for larger regurgitant volumes in larger individuals. As shown in Figure 2, the distributions of TR grades by TR fraction vs indexed TR volume are strikingly similar.

Current guidelines recommend using CMR in the evaluation of right heart size and function in patients with severe TR and the assessment of reverse RV remodeling (both atrial and ventricular)^33^. An accurate TR classification is important because functional TR subtypes follow different clinical courses. Gavazzoni et al. showed a 2.15-fold higher risk of a 1-year composite of death and hospitalization in patients with secondary ventricular TR compared to secondary atrial TR^34^, emphasizing the importance of early diagnosis of right ventricular dysfunction. Several studies correlated different 2D flow CMR parameters of RV volume and function with increased all-cause mortality in TR patients; this includes RVEDV, RVESV, RV ejection fraction and RV strain (Table 4)^13–15^. 4D flow RV assessment can be useful in patients with pulmonary hypertension and RV diastolic dysfunction, which frequently coexist^35,36^. In our study, different echocardiographic grades were not fully distinguishable by RVEDV indexed by BSA. We also found a moderate correlation between indexed RVEDV and TR volume as well as indexed RVEDV and TR fraction. This highlights the need for more routine CMR evaluation of the RV in TR patients, as TR severity may not always indicate the extent of RV dilation.

**Table 4.**
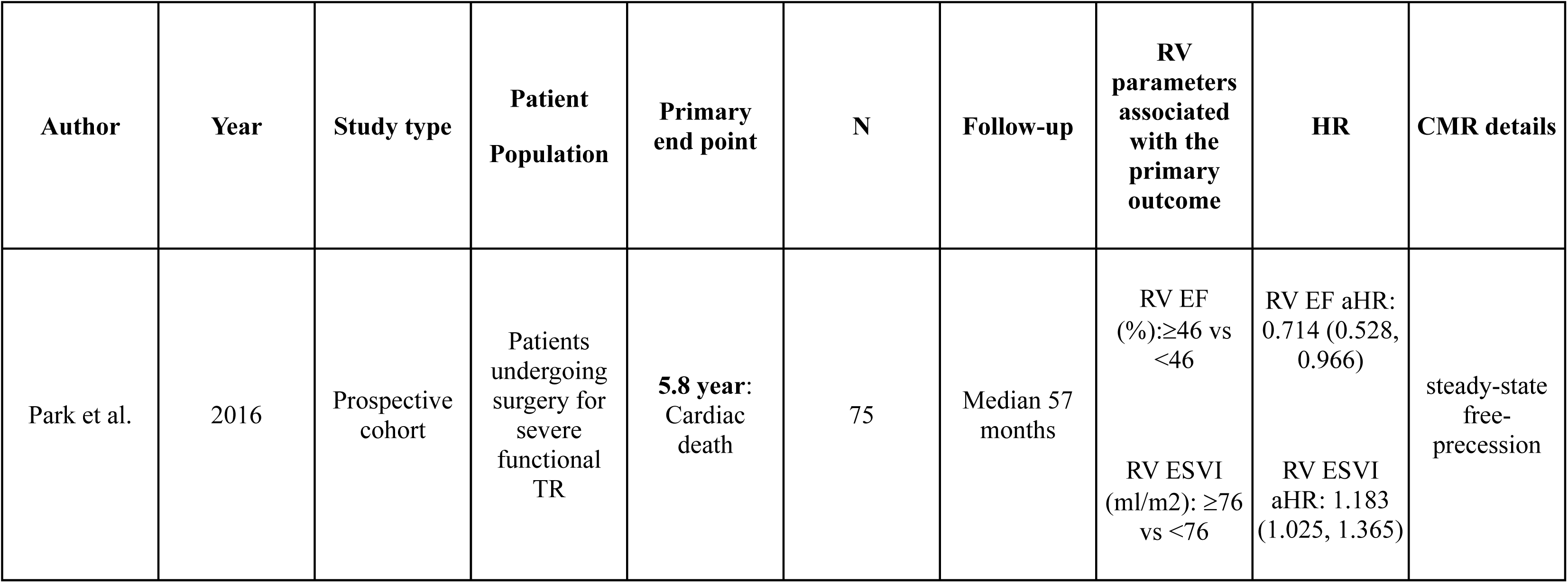
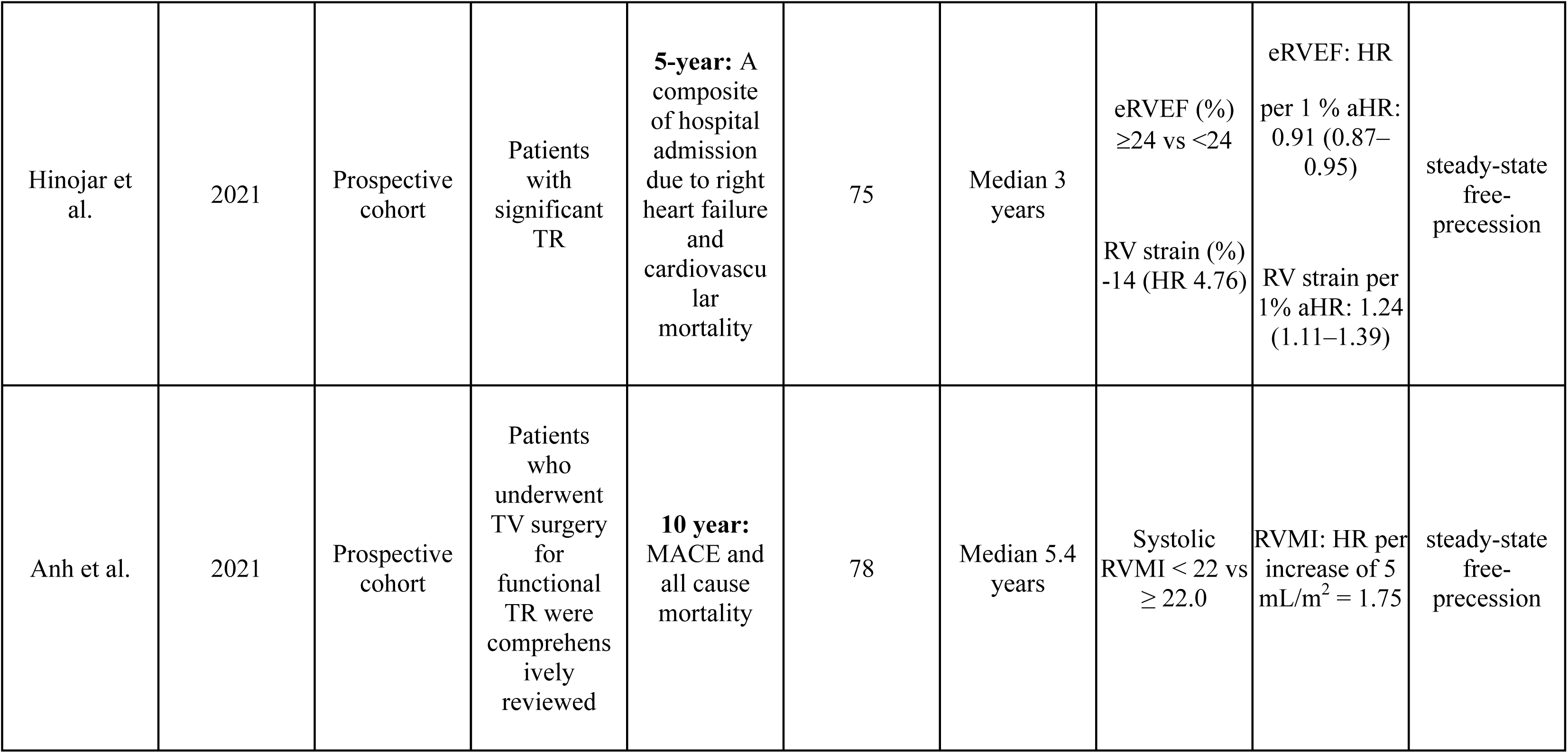
Cardiac MRI (CMR) thresholds for right ventricular (RV) indices proposed in selected prospective studies of tricuspid regurgitation (TR) patients. Abbreviations: HR: hazard ratio; aHR: adjusted hazard ratio; EF: ejection fraction; RVESVI: right ventricular end-systolic volume index; RVMI: right ventricular motion index; MACE: major adverse cardiovascular events; TV: tricuspid valve.

| Author | Year | Study type | Patient Population | Primary end point | N | Follow-up | RV parameters associated with the primary outcome | HR | CMR details |
| --- | --- | --- | --- | --- | --- | --- | --- | --- | --- |
| Park et al. | 2016 | Prospective cohort | Patients undergoing surgery for severe functional TR | 5.8 year: Cardiac death | 75 | Median 57 months | RV EF (%): ≥46 vs <46<br>RV ESVI (ml/m2): ≥76 vs <76 | RV EF aHR: 0.714 (0.528, 0.966)<br>RV ESVI aHR: 1.183 (1.025, 1.365) | steady-state free-precession |
| Hinojar et al. | 2021 | Prospective cohort | Patients with significant TR | **5-year:** A composite of hospital admission due to right heart failure and cardiovascular mortality | 75 | Median 3 years | eRVEF (%)<br>≥24 vs <24<br>RV strain (%)<br>-14 (HR 4.76) | eRVEF: HR per 1 % aHR: 0.91 (0.87–0.95)<br>RV strain per 1% aHR: 1.24 (1.11–1.39) | steady-state free-precession |
| Anh et al. | 2021 | Prospective cohort | Patients who underwent TV surgery for functional TR were comprehensively reviewed | **10 year:** MACE and all cause mortality | 78 | Median 5.4 years | Systolic RVMI < 22 vs ≥ 22.0 | RVMI: HR per increase of 5 mL/m<sup>2</sup> = 1.75 | steady-state free-precession |

The results of our study raise several important questions: What is the ‘gold standard’ for grading TR. Is it TTE, CMR, or other modalities? Which parameter is most accurate (TR volume, TR fraction or others)? What is the impact of dynamic nature of TR? Should we consider stress testing or incorporating invasive hemodynamic assessment in the evaluation of these patients? What is the impact of extra-valve cardiac abnormalities (RV size and function, PA pressures, RV-PA coupling) on the grading of TR? Should multi-modality imaging be considered in patients referred for the newly commercially available TV repair and replacement technologies^37^?. Will patients who have concordantly severe TR on TTE and CMR derive the most benefit from TR interventions? These questions require further collaborative investigations and will inform the quickly growing field of transcatheter tricuspid valve interventions.

### Limitations

Our pilot study has several limitations: First, TR severity is preload-dependent and therefore affected by volume status and diuretics. As such, the discrepancies observed in some patients might be explained by the time interval between the TTE and CMR imaging. This was unfortunately unavoidable considering the logistics of securing a research 4D-CMR for patients who are largely travelling to our teritiary center. However, we observed no significant changes in body weight or diuretic dose between the two scans suggesting that this difference is not predominantly related to difference in loading conditions. Second, some patients had atrial fibrillation, which may have introduced beat-to-beat variability and can impact our assessment.

Third, patients with pacemakers or ICDs were excluded to minimize imaging artifacts. Therefore, our findings may not be fully generalizable to this cohort. Fourth, determining the ideal imaging modality to assess TR require benchnakring TR grades between modalities against long-term clinical outcomes. This necessitates a larger study which was beyond the scope of this exploratory study.

### Conclusions

Grading of tricuspid regurigation is frequently discordant between TEE and CMR. Further studies are needed to elucidate the clinical impact of concordant/discordant TR grading on multi-modality imaging.

## Data Availability

Anonymized data presented in the manuscript can be made available upon request.

---

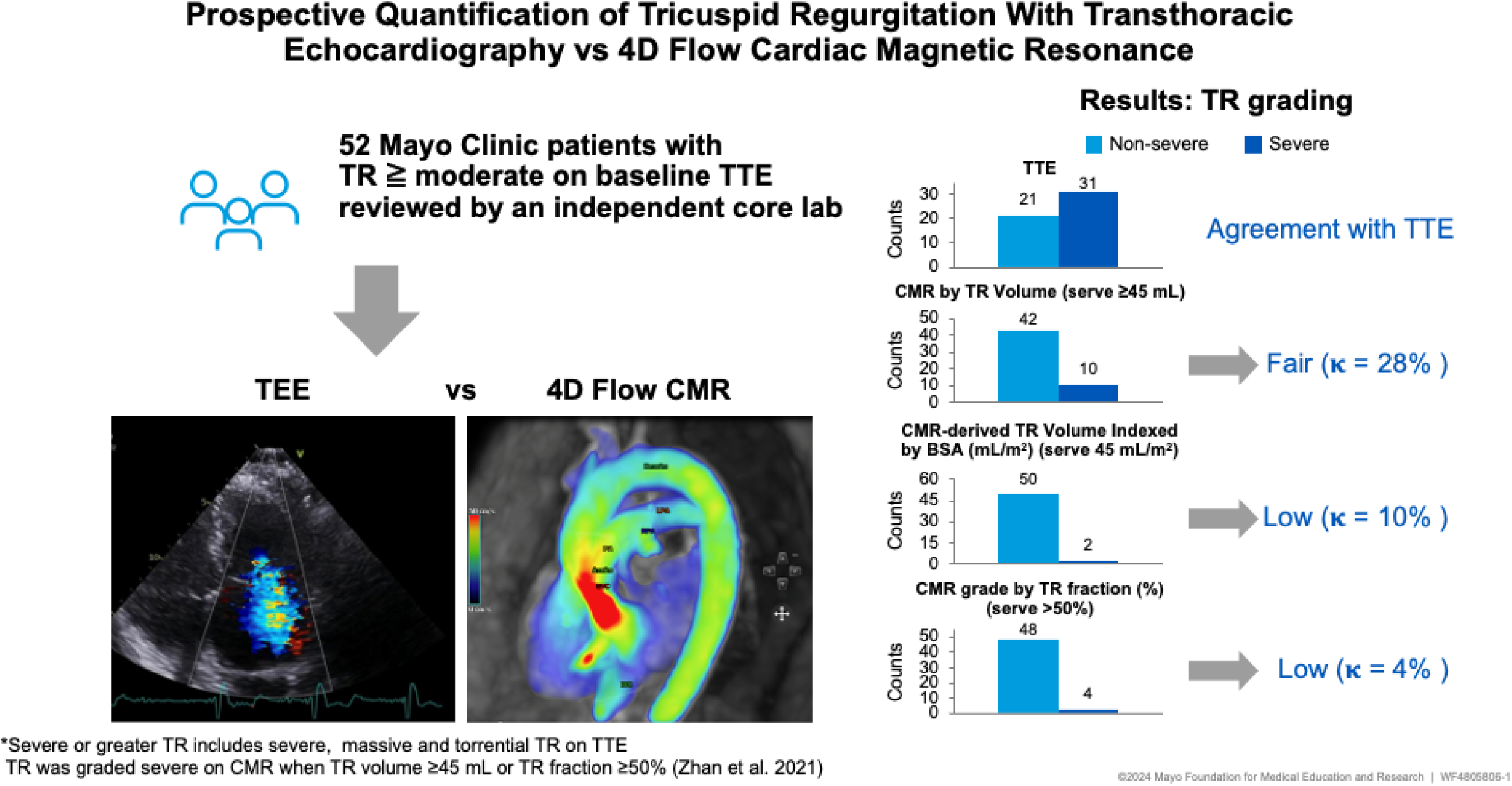

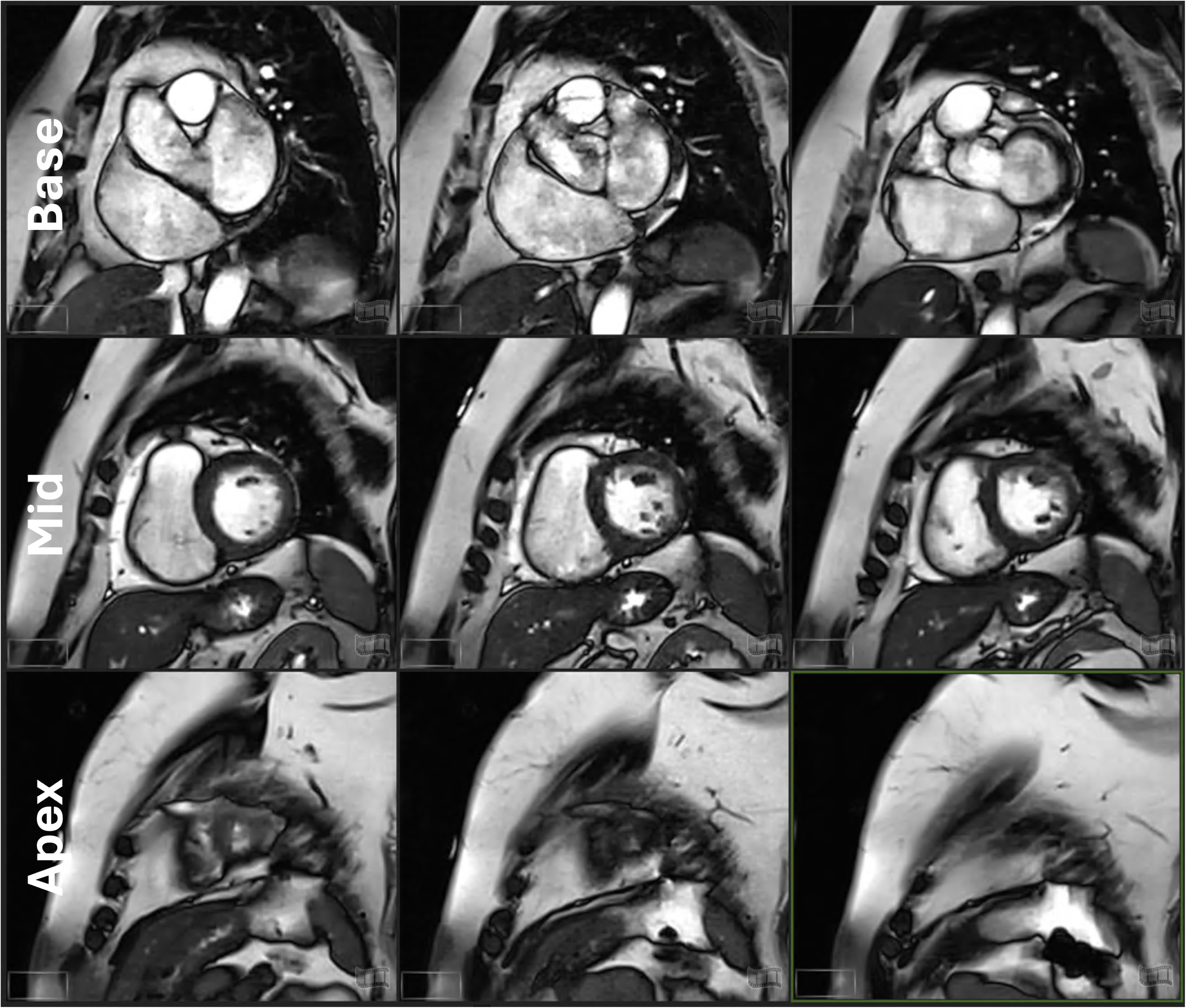

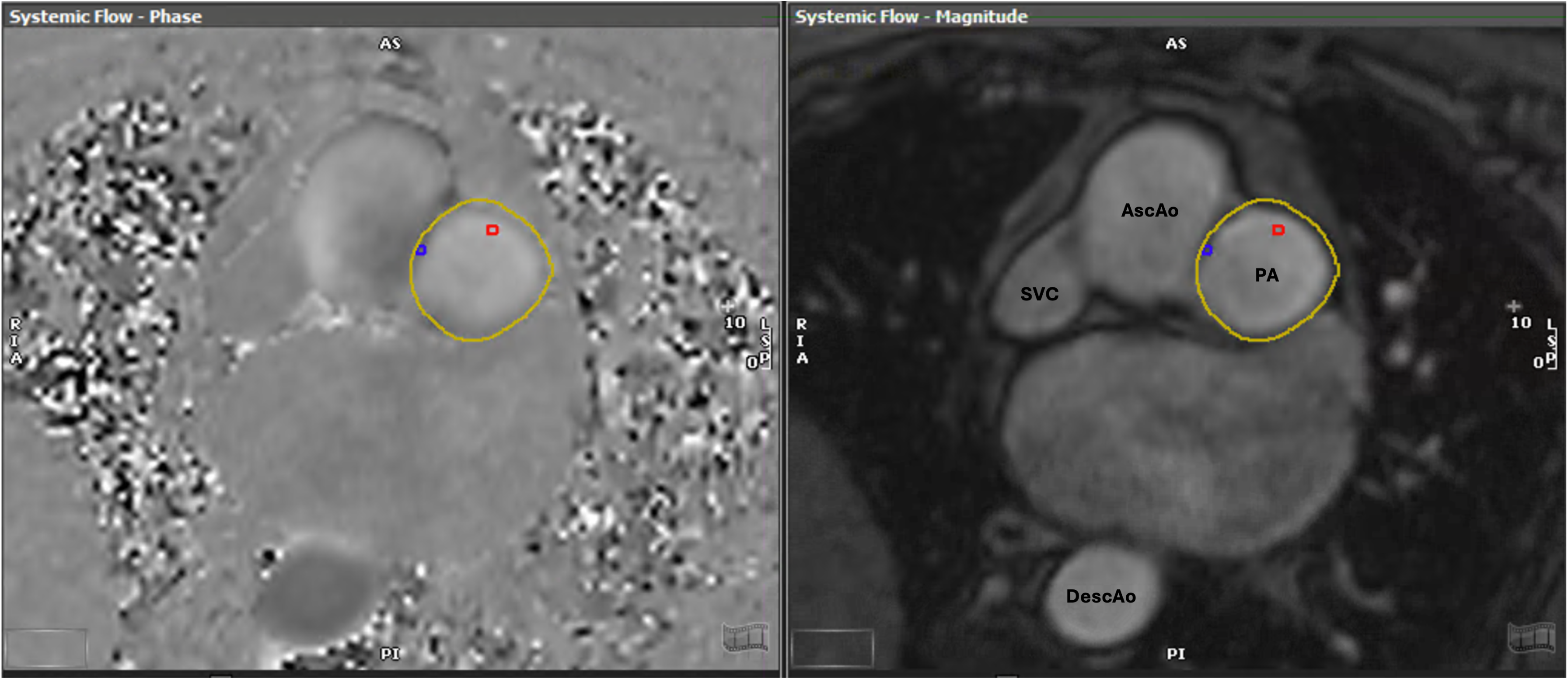

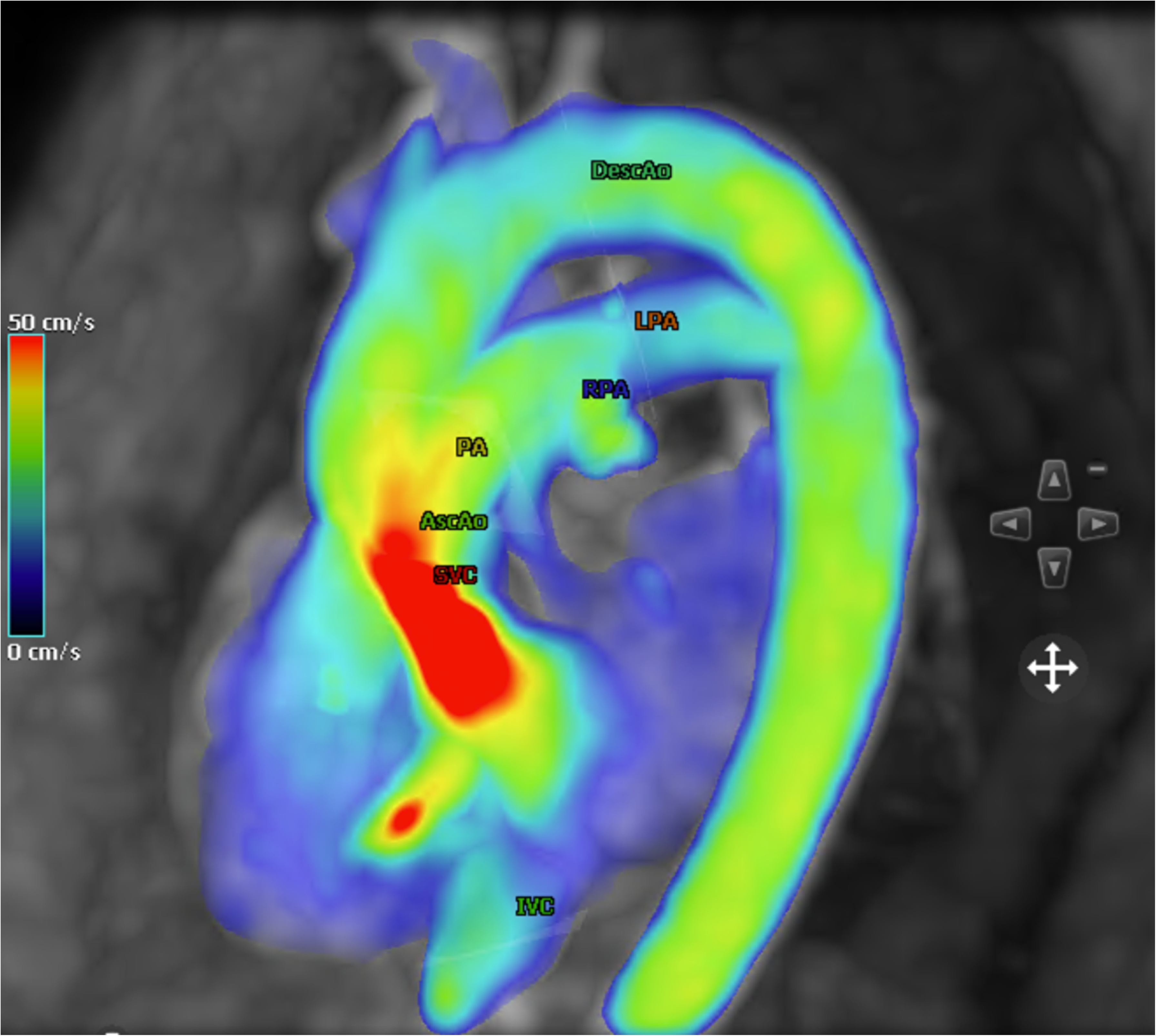

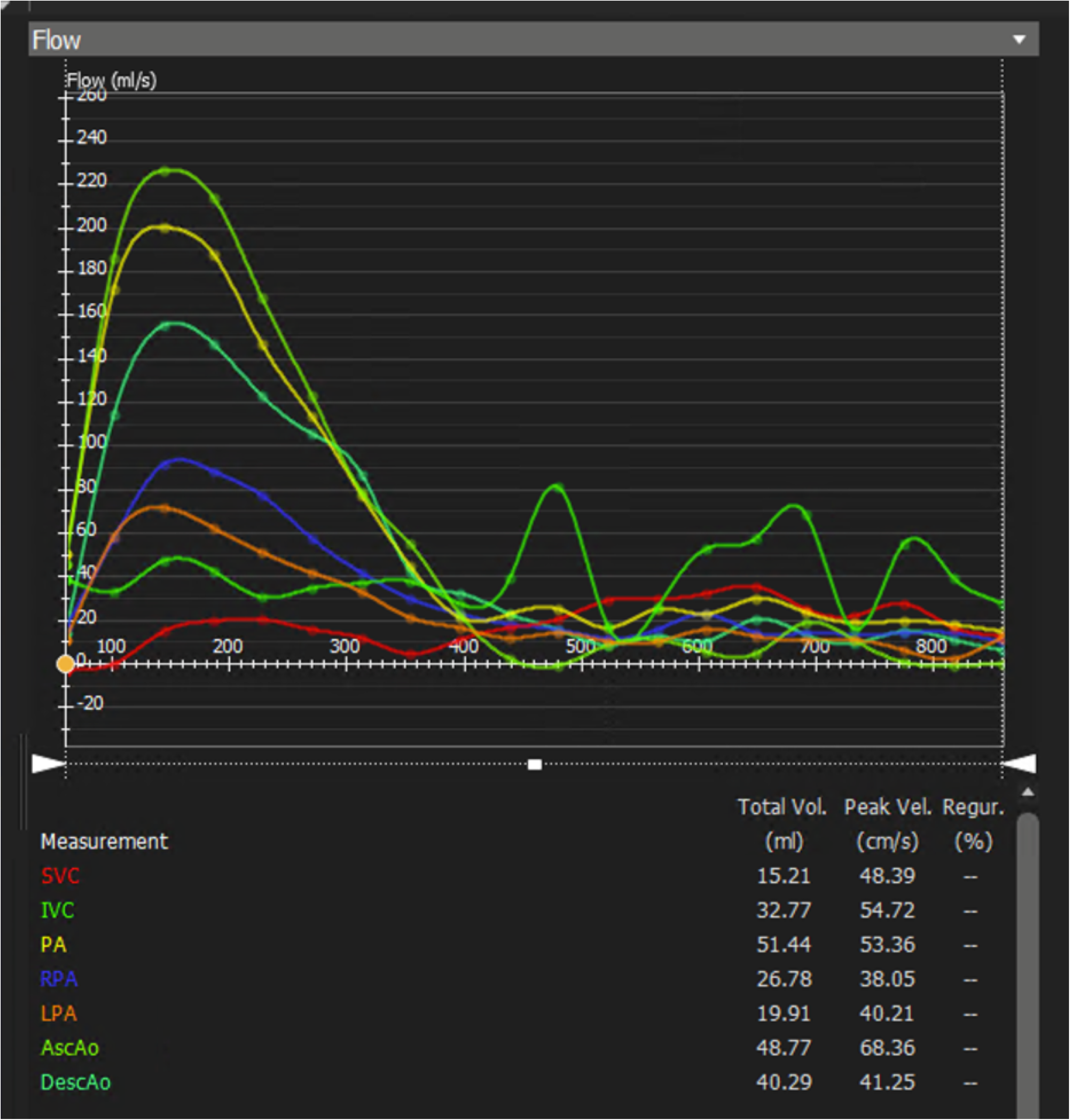

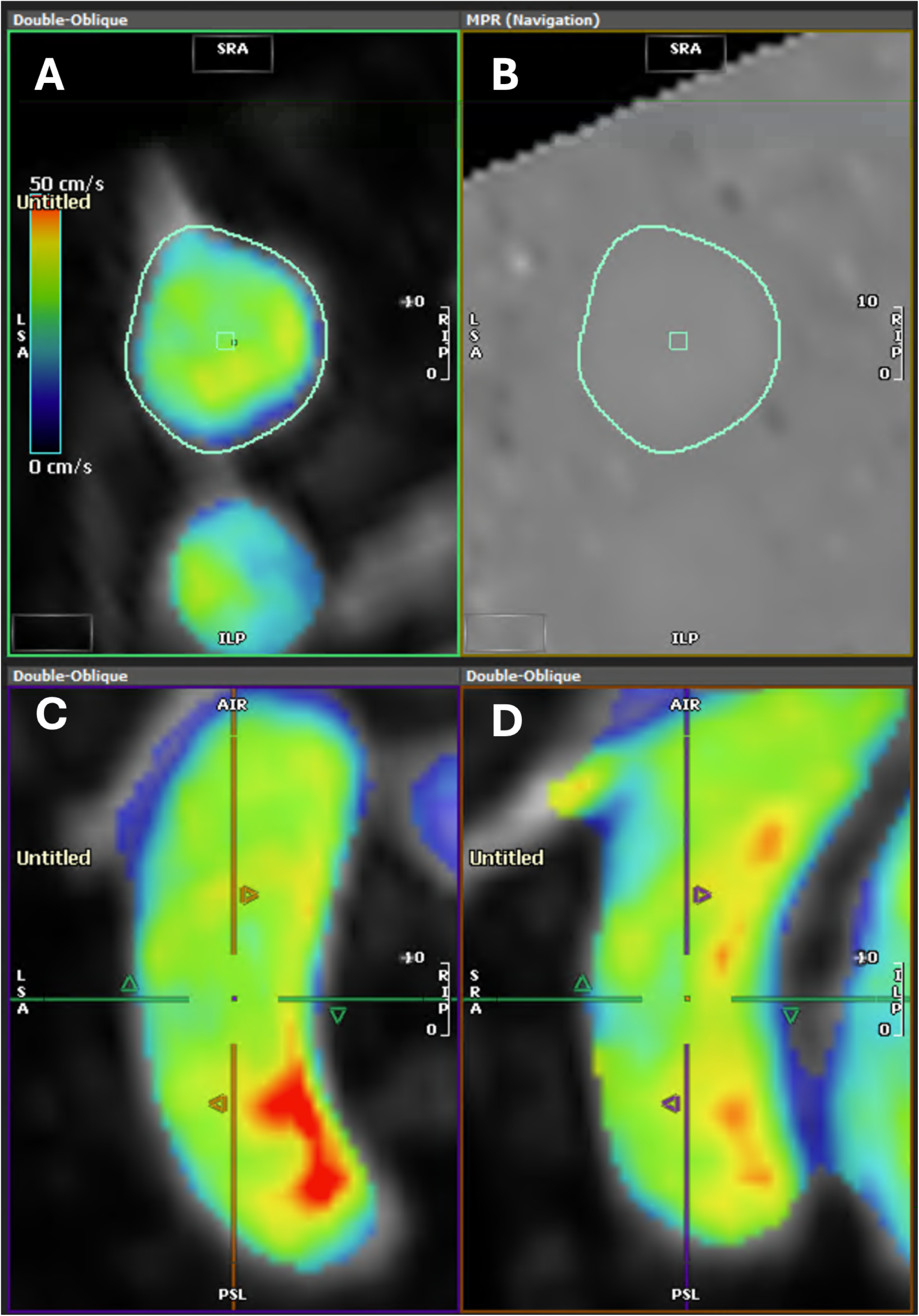

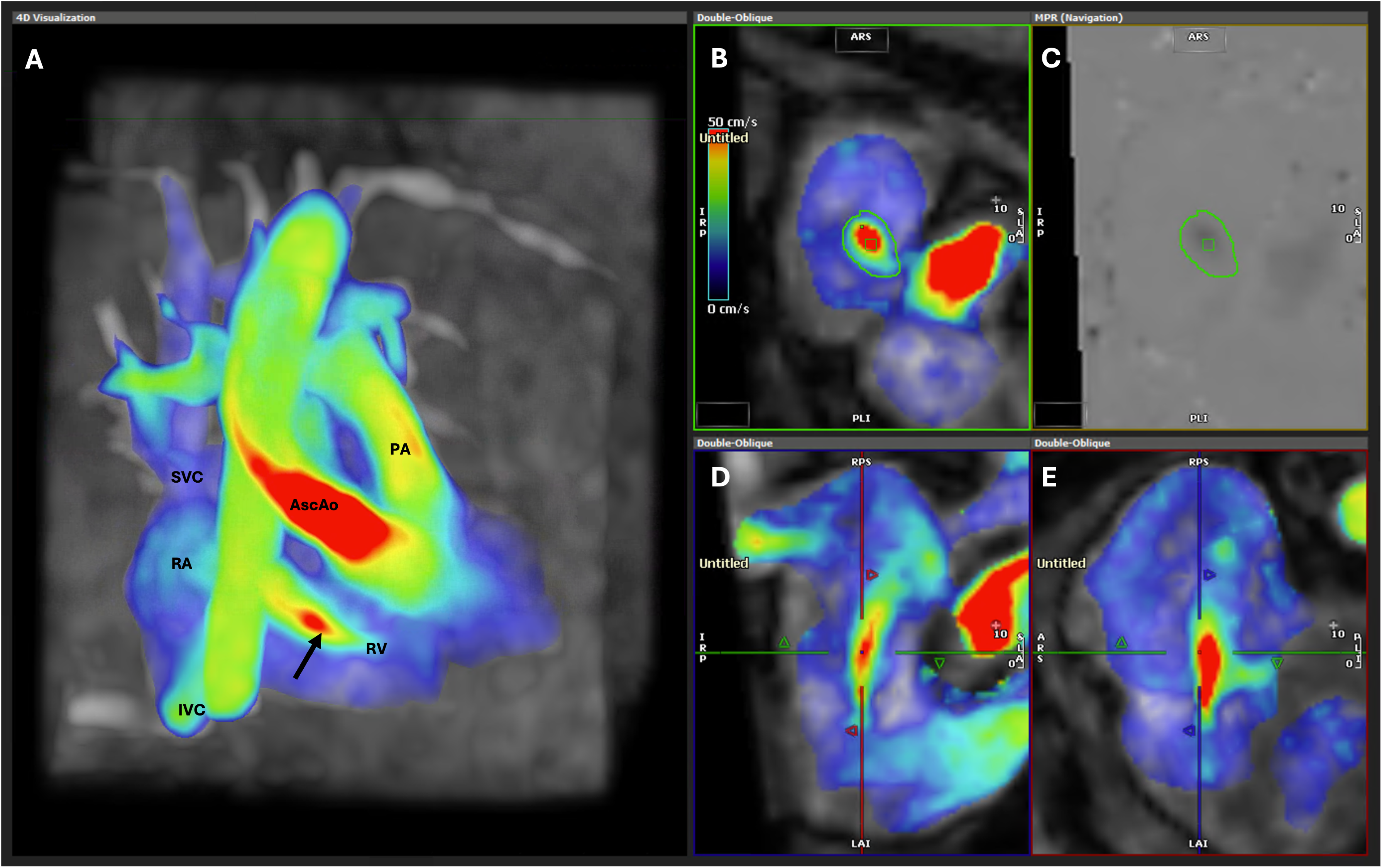

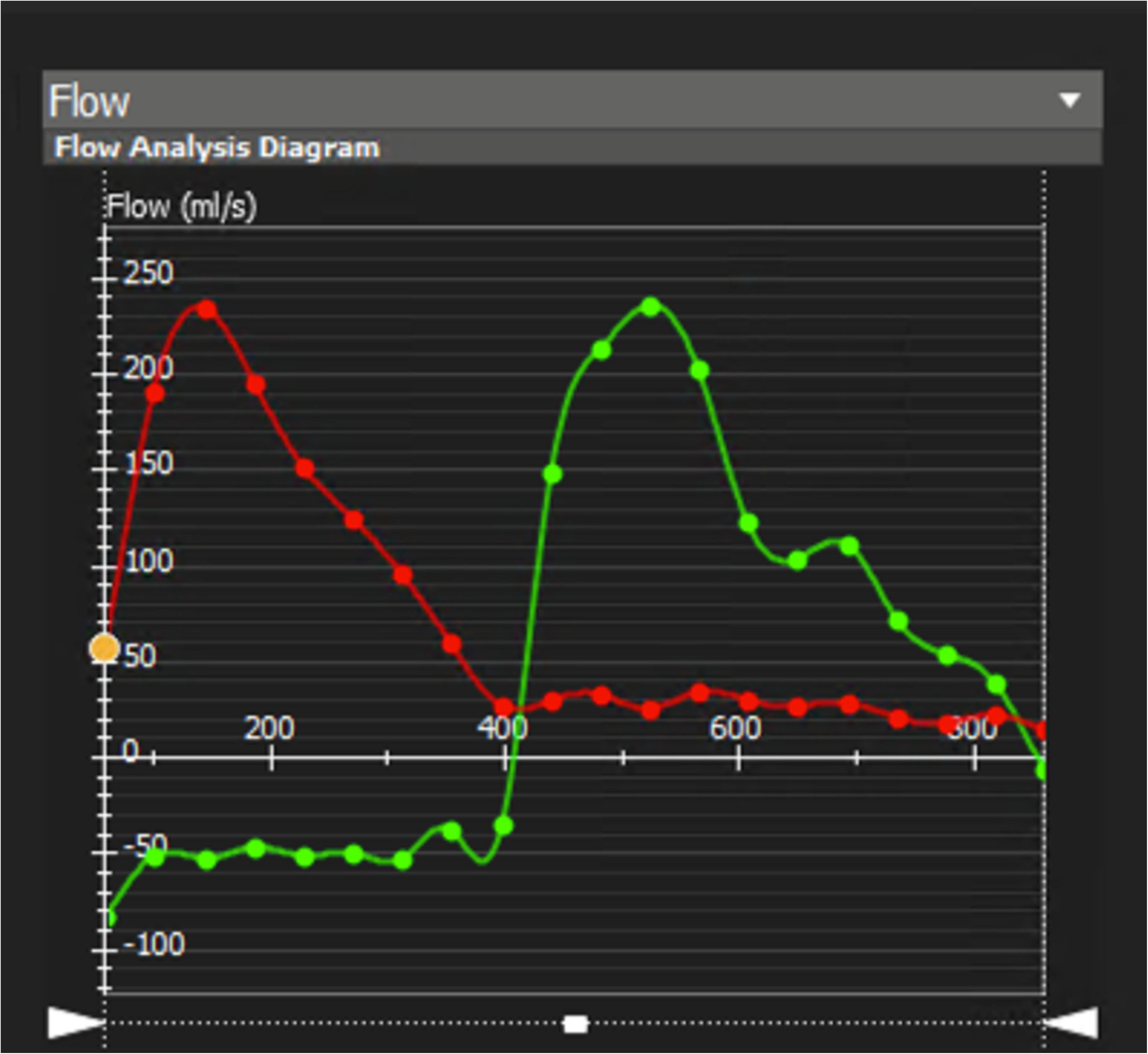

## References

1. Hahn RT. Tricuspid Regurgitation. N Engl J Med. 2023;388:1876–1891. doi: 10.1056/NEJMra2216709

2. Alqahtani F, Berzingi CO, Aljohani S, Hijazi M, Al-Hallak A, Alkhouli M. Contemporary Trends in the Use and Outcomes of Surgical Treatment of Tricuspid Regurgitation. J Am Heart Assoc. 2017;6. doi: 10.1161/JAHA.117.007597

3. Otto CM, Nishimura RA, Bonow RO, Carabello BA, Erwin JP, 3rd, Gentile F, Jneid H, Krieger EV, Mack M, McLeod C, et al. 2020 ACC/AHA Guideline for the Management of Patients With Valvular Heart Disease: Executive Summary: A Report of the American College of Cardiology/American Heart Association Joint Committee on Clinical Practice Guidelines. Circulation. 2021;143:e35–e71. doi: 10.1161/CIR.0000000000000932

4. Alkhouli M, Lopez JJ, Mathew V. Transcatheter Therapy for Severe Tricuspid Regurgitation: Learning to Understand the Forgotten Valve. J Am Coll Cardiol. 2019;74:3009–3012. doi: 10.1016/j.jacc.2019.09.029

5. Grapsa J, Praz F, Sorajja P, Cavalcante JL, Sitges M, Taramasso M, Piazza N, Messika-Zeitoun D, Michelena HI, Hamid N, et al. Tricuspid Regurgitation: From Imaging to Clinical Trials to Resolving the Unmet Need for Treatment. JACC Cardiovasc Imaging. 2024;17:79–95. doi: 10.1016/j.jcmg.2023.08.013

6. Kawsara A, Alqahtani F, Nkomo VT, Eleid MF, Pislaru SV, Rihal CS, Nishimura RA, Schaff HV, Crestanello JA, Alkhouli M. Determinants of Morbidity and Mortality Associated With Isolated Tricuspid Valve Surgery. J Am Heart Assoc. 2021;10:e018417. doi: 10.1161/JAHA.120.018417

7. Lara-Breitinger KM, Scott CG, Nkomo VT, Pellikka PA, Kane GC, Chaliki HP, Shapiro BP, Eleid MF, Alkhouli M, Greason KL, et al. Tricuspid Regurgitation Impact on Outcomes (TRIO): A Simple Clinical Risk Score. Mayo Clin Proc. 2022;97:1449–1461. doi: 10.1016/j.mayocp.2022.05.015

8. Welle GA, Hahn RT, Lindenfeld J, Lin G, Nkomo VT, Hausleiter J, Lurz PC, Pislaru SV, Davidson CJ, Eleid MF. New Approaches to Assessment and Management of Tricuspid Regurgitation Before Intervention. JACC Cardiovasc Interv. 2024;17:837–858. doi: 10.1016/j.jcin.2024.02.034

9. Hahn RT, Badano LP, Bartko PE, Muraru D, Maisano F, Zamorano JL, Donal E. Tricuspid regurgitation: recent advances in understanding pathophysiology, severity grading and outcome. Eur Heart J Cardiovasc Imaging. 2022;23:913–929. doi: 10.1093/ehjci/jeac009

10. Hahn RT, Weckbach LT, Noack T, Hamid N, Kitamura M, Bae R, Lurz P, Kodali SK, Sorajja P, Hausleiter J, et al. Proposal for a Standard Echocardiographic Tricuspid Valve Nomenclature. JACC Cardiovasc Imaging. 2021;14:1299–1305. doi: 10.1016/j.jcmg.2021.01.012

11. Weckbach LT, Orban M, Kitamura M, Hamid N, Lurz P, Hahn RT, Sorajja P, Nabauer M, Noack T, Hausleiter J. Tricuspid Valve Morphology and Outcome in Patients Undergoing Transcatheter Tricuspid Valve Edge-to-Edge Repair. JACC Cardiovasc Interv. 2022;15:567–569. doi: 10.1016/j.jcin.2021.12.028

12. Maffessanti F, Gripari P, Pontone G, Andreini D, Bertella E, Mushtaq S, Tamborini G, Fusini L, Pepi M, Caiani EG. Three-dimensional dynamic assessment of tricuspid and mitral annuli using cardiovascular magnetic resonance. Eur Heart J Cardiovasc Imaging. 2013;14:986–995. doi: 10.1093/ehjci/jet004

13. Park JB, Kim HK, Jung JH, Klem I, Yoon YE, Lee SP, Park EA, Hwang HY, Lee W, Kim KH, et al. Prognostic Value of Cardiac MR Imaging for Preoperative Assessment of Patients with Severe Functional Tricuspid Regurgitation. Radiology. 2016;280:723–734. doi: 10.1148/radiol.2016151556

14. Hinojar R, Gomez AG, Garcia-Martin A, Monteagudo JM, Fernandez-Mendez MA, de Vicente AG, Salinas GLA, Zamorano JL, Fernandez-Golfin C. Impact of right ventricular systolic function in patients with significant tricuspid regurgitation. A cardiac magnetic resonance study. Int J Cardiol. 2021;339:120–127. doi: 10.1016/j.ijcard.2021.07.023

15. Ahn Y, Koo HJ, Kang JW, Choi WJ, Kim DH, Song JM, Kang DH, Song JK, Kim JB, Jung SH, et al. Prognostic Implication of Right Ventricle Parameters Measured on Preoperative Cardiac MRI in Patients with Functional Tricuspid Regurgitation. Korean J Radiol. 2021;22:1253–1265. doi: 10.3348/kjr.2020.1084

16. Driessen MMP, Schings MA, Sieswerda GT, Doevendans PA, Hulzebos EH, Post MC, Snijder RJ, Westenberg JJM, van Dijk APJ, Meijboom FJ, et al. Tricuspid flow and regurgitation in congenital heart disease and pulmonary hypertension: comparison of 4D flow cardiovascular magnetic resonance and echocardiography. J Cardiovasc Magn Reson. 2018;20:5. doi: 10.1186/s12968-017-0426-7

17. Feneis JF, Kyubwa E, Atianzar K, Cheng JY, Alley MT, Vasanawala SS, Demaria AN, Hsiao A. 4D flow MRI quantification of mitral and tricuspid regurgitation: Reproducibility and consistency relative to conventional MRI. J Magn Reson Imaging. 2018;48:1147–1158. doi: 10.1002/jmri.26040

18. Zhan Y, Senapati A, Vejpongsa P, Xu J, Shah DJ, Nagueh SF. Comparison of Echocardiographic Assessment of Tricuspid Regurgitation Against Cardiovascular Magnetic Resonance. JACC Cardiovasc Imaging. 2020;13:1461–1471. doi: 10.1016/j.jcmg.2020.01.008

19. Medvedofsky D, Leon Jimenez J, Addetia K, Singh A, Lang RM, Mor-Avi V, Patel AR. Multi-parametric quantification of tricuspid regurgitation using cardiovascular magnetic resonance: A comparison to echocardiography. Eur J Radiol. 2017;86:213–220. doi: 10.1016/j.ejrad.2016.11.025

20. Spiewak M, Klopotowski M, Kowalik E, Mazurkiewicz L, Kozuch K, Petryka-Mazurkiewicz J, Milosz-Wieczorek B, Witkowski A, Klisiewicz A, Marczak M. Comparison of mitral regurgitation severity assessments based on magnetic resonance imaging and echocardiography in patients with hypertrophic cardiomyopathy. Sci Rep. 2021;11:19902. doi: 10.1038/s41598-021-99446-y

21. Krieger EV, Lee J, Branch KR, Hamilton-Craig C. Quantitation of mitral regurgitation with cardiac magnetic resonance imaging: a systematic review. Heart. 2016;102:1864–1870. doi: 10.1136/heartjnl-2015-309054

22. Uretsky S, Gillam L, Lang R, Chaudhry FA, Argulian E, Supariwala A, Gurram S, Jain K, Subero M, Jang JJ, et al. Discordance Between Echocardiography and MRI in the Assessment of Mitral Regurgitation Severity. Journal of the American College of Cardiology. 2015;65:1078–1088. doi: doi:10.1016/j.jacc.2014.12.047

23. Lopez-Mattei JC, Ibrahim H, Shaikh KA, Little SH, Shah DJ, Maragiannis D, Zoghbi WA. Comparative Assessment of Mitral Regurgitation Severity by Transthoracic Echocardiography and Cardiac Magnetic Resonance Using an Integrative and Quantitative Approach. The American Journal of Cardiology. 2016;117:264–270. doi: 10.1016/j.amjcard.2015.10.045

24. Penicka M, Vecera J, Mirica DC, Kotrc M, Kockova R, Van Camp G. Prognostic implications of magnetic resonance–derived quantification in asymptomatic patients with organic mitral regurgitation: comparison with Doppler echocardiography–derived integrative approach. Circulation. 2018;137:1349–1360.

25. Zhan Y, Debs D, Khan MA, Nguyen DT, Graviss EA, Khalaf S, Little SH, Reardon MJ, Nagueh S, Quinones MA, et al. Natural History of Functional Tricuspid Regurgitation Quantified by Cardiovascular Magnetic Resonance. J Am Coll Cardiol. 2020;76:1291–1301. doi: 10.1016/j.jacc.2020.07.036

26. Zoghbi WA, Adams D, Bonow RO, Enriquez-Sarano M, Foster E, Grayburn PA, Hahn RT, Han Y, Hung J, Lang RM, et al. Recommendations for Noninvasive Evaluation of Native Valvular Regurgitation: A Report from the American Society of Echocardiography Developed in Collaboration with the Society for Cardiovascular Magnetic Resonance. J Am Soc Echocardiogr. 2017;30:303–371. doi: 10.1016/j.echo.2017.01.007

27. Topilsky Y, Bae R, Nkomo V, Gavazzoni M. An Integrative, Multiparametric Approach to Tricuspid Regurgitation Evaluation: A Case-Based Illustration. JACC Case Rep. 2023;25:102050. doi: 10.1016/j.jaccas.2023.102050

28. Wang TKM, Reyaldeen R, Akyuz K, Popovic ZB, Gillinov AM, Xu B, Griffin BP, Desai MY. Echocardiography Versus Magnetic Resonance Imaging Quantification and Novel Algorithm for Isolated Severe Tricuspid Regurgitation. Am J Cardiol. 2023;211:40–48. doi: 10.1016/j.amjcard.2023.10.062

29. Renella P, Aboulhosn J, Lohan DG, Jonnala P, Finn JP, Satou GM, Williams RJ, Child JS. Two-dimensional and Doppler echocardiography reliably predict severe pulmonary regurgitation as quantified by cardiac magnetic resonance. J Am Soc Echocardiogr. 2010;23:880–886. doi: 10.1016/j.echo.2010.05.019

30. Hahn RT. State-of-the-Art Review of Echocardiographic Imaging in the Evaluation and Treatment of Functional Tricuspid Regurgitation. Circ Cardiovasc Imaging. 2016;9. doi: 10.1161/CIRCIMAGING.116.005332

31. Wang TKM, Unai S, Xu B. Contemporary review in the multi-modality imaging evaluation and management of tricuspid regurgitation. Cardiovasc Diagn Ther. 2021;11:804–817. doi: 10.21037/cdt.2020.01.06

32. Seo J, Hong YJ, Batbayar U, Kim DY, Cho I, Kim YJ, Hong GR, Ha JW, Shim CY. Prognostic value of functional tricuspid regurgitation quantified by cardiac magnetic resonance in heart failure. Eur Heart J Cardiovasc Imaging. 2023;24:742–750. doi: 10.1093/ehjci/jeac224

33. Correction to: 2020 ACC/AHA Guideline for the Management of Patients With Valvular Heart Disease: A Report of the American College of Cardiology/American Heart Association Joint Committee on Clinical Practice Guidelines. Circulation. 2023;148:e8. doi: 10.1161/CIR.0000000000001177

34. Gavazzoni M, Heilbron F, Badano LP, Radu N, Cascella A, Tomaselli M, Perelli F, Caravita S, Baratto C, Parati G, et al. Corrigendum: The atrial secondary tricuspid regurgitation is associated to more favorable outcome than the ventricular phenotype. Front Cardiovasc Med. 2023;10:1169907. doi: 10.3389/fcvm.2023.1169907

35. Cain MT, Schafer M, Ross LK, Ivy DD, Mitchell MB, Fenster BE, Bull TM, Barker AJ, Vargas D, Hoffman JRH. 4D-Flow MRI intracardiac flow analysis considering different subtypes of pulmonary hypertension. Pulm Circ. 2023;13:e12307. doi: 10.1002/pul2.12307

36. Barker N, Fidock B, Johns CS, Kaur H, Archer G, Rajaram S, Hill C, Thomas S, Karunasaagarar K, Capener D, et al. A Systematic Review of Right Ventricular Diastolic Assessment by 4D Flow CMR. Biomed Res Int. 2019;2019:6074984. doi: 10.1155/2019/6074984

37. Kodali S, Hahn RT, Makkar R, Makar M, Davidson CJ, Puthumana JJ, Zahr F, Chadderdon S, Fam N, Ong G, et al. Transfemoral tricuspid valve replacement and one-year outcomes: the TRISCEND study. Eur Heart J. 2023;44:4862–4873. doi: 10.1093/eurheartj/ehad667

